# Body surface colonic mapping detects meal and bisacodyl-induced colonic motility in patients with chronic constipation

**DOI:** 10.64898/2026.02.08.26345865

**Authors:** Annelies Verheyden, Phil G. Dinning, Greg O’Grady, Jan Tack, Jonathan C. Erickson

## Abstract

Chronic constipation is highly prevalent, and cases refractory to treatment are particularly challenging to manage. High-resolution colonic manometry (HRM) is used to further evaluate these patients to identify cases of intrinsic motor dysfunction (underlying myopathy or neuropathy). However, HRM is invasive and resource-intensive, limiting uptake and clinical utility. This study presents Body Surface Colonic Mapping (BSCM), a non-invasive cutaneous electrical recording technique, as a clinical alternative. Simultaneous recordings from HRM (36-channel) and BSCM (8×8 electrode array) were performed in 10 patients with chronic refractory constipation. Lower gut symptom scores were also tracked patients over the duration of the recording. Motility was assessed during meal and bisacodyl challenges. We optimized BSCM signal processing specifically to detect high-amplitude propagating contractions (HAPCs) evoked by bisacodyl. Analysis included time-frequency quantification of motility indices and blinded visual assessment by domain experts to classify the presence or absence of motor responses. BSCM motility indices showed strong correlation with HRM for both meal (*r* = 0.86) and bisacodyl (*r* = 0.69) responses. Expert visual analysis yielded concordant classification between BSCM and HRM in the majority (87.5 ± 9.6%) of cases. Furthermore, BSCM identified distinct, patient-specific symptom-motility associations during the meal response. BSCM accurately detects meal- and stimulant-induced increases in colonic motility with high fidelity to invasive HRM. As a non-invasive method that is easy to apply with minimal resource and time requirements, BSCM is well-positioned for clinical translation as a scalable diagnostic tool to elucidate symptom-motility associations and guide personalized management in refractory chronic constipation.

## INTRODUCTION

Chronic constipation affects more than 10% of the population *(1, 2)* and can significantly impair quality of life *(3)*, yet its pathophysiology often remains elusive. For patients refractory to standard treatments, high-resolution colonic manometry (HRM) can be used to evaluate neuromuscular integrity and characterize meal and pharmacological responses.

Typically, HRM protocols include a meal *(4)* and occasionally an intraluminal infusion of a laxative such as bisacodyl *(5)*. In health the meal induces a robust rise in cyclic motor patterns *(6)* whilst bisacodyl induces a series of the propulsive high amplitude propagating contractions (HAPCs) *(7, 8)*. In patients with severe constipation both the meal and bisacodyl response can be attenuated, which may indicate a myogenic and or neurogenic disorder *(7, 9, 10)*. Despite its potential diagnostic value, HRM remains limited to a small number of specialty centers worldwide. The technique requires some form of bowel preparation, colonoscopic catheter placement and a prolonged day-long stay in the clinical center. Analysis of the acquired data tends to be center-specific and time-intensive, factors that further restrict its widespread clinical use. This highlights the need for a non-invasive, easy-to-apply, and standardized alternative for assessing colonic motor responses.

Body-surface gastric mapping, a non-invasive technology for measuring gastric myoelectrical activity, has recently gained international regulatory approval for assessing gastric motility disorders. The same technology can be applied to the colon. Recently, body-surface colonic mapping (BSCM) has been shown, via validation against HRM, to detect a postprandial increase in the cyclic motor pattern in a small cohort of healthy controls *(11)*. However, it has not been applied to study the meal responses in patients with function gut disorders. Moreover, the BSCM technique has not yet been applied or validated for detection of bisacodyl-induced colonic motility, which typically consists of a sequence of HAPCs.

In this study, we made simultaneous recordings of HRM and BSCM, with the aim of identifying colonic responses to a meal and intraluminal bisacodyl stimuli in patients with chronic constipation. Herein we present the key advance that BSCM reliably detects colonic motility stimulated by bisacodyl, primarily clusters of HAPCs. We also show that BSCM reliably identifies meal response activity in this patient cohort, extending the findings previously obtained in a small cohort of healthy controls *(11)*. Additionally, BSCM successfully correlates patient-reported symptoms with increased motility, offering insight into the potential mechanisms of symptom genesis in chronic constipation.

## RESULTS

### Patient demographics and study characteristics

Twelve patients (3M/9F, mean age 49 years, (range 27-67), mean BMI 21.2 kg/m^2^ (range 16.7-33.8 kg/m^2^) completed the study. Ten were included for full analysis (3M/7F; 48.2 years, range 27-67; mean BMI 21.1kg/m^2^). One was excluded because the BSCM recording terminated prior to a delayed bisacodyl infusion, and the other on account of uncharacteristic very low signal-to-noise-ratio BSCM signals likely due to an undetected reference electrode disconnection early in the study. Of the 10 studies carried forward for full analysis, two BSCM arrays were used on 3 patients while a single array was used on the remaining 7 patients to achieve sufficient abdominal coverage. The catheter tip was clipped to the ascending colon in 6 patients and to the hepatic flexure for the remaining 4. The study duration was 218 ± 44 min, range=173-315 min.

### HRM manual identification and characterization of bisacodyl responses

Post-bisacodyl HAPCs were identified by manual analysis in 9 out of 10 patients. In these 9 patients 69 HAPC were identified (7.7±3.8 HAPC/patient; range, 2-15). The remaining patient responded to bisacodyl with a non-HAPCs, high-amplitude, uncoordinated rhythmic motility pattern with dominant frequencies of 4.0 and 12.3 cpm spatially confined to the rectosigmoid region.

The response time from bisacodyl infusion to the first HAPCs event was 7.3 ± 3.8 min (1.3 -13.6 min). The response duration (time elapsed between first to last HAPCs) was 31.8 ± 19.6 min (2.96 - 51.5 min). The distribution of event intervals (= *T*_*HAPC*_[*i* + 1] − *T*_*HAPC*_[*i*]) was concentrated in the 1 - 4 minute range (median ± MADM = 2.7 ± 1.2 min, 80% < 5 min). This equates to event rates (=1/interval) of 0.37 ± 0.18 cpm with 71% in the range of 0.25 - 1 cpm (see also Supplementary Materials fig. S4).

### Concordance of BSCM and HRM Motility Indices

A comparison of motility index (MI) and spectrograms from HRM and BSCM during meal and bisacodyl response epochs is shown for one patient in Fig. 2. Both BSCM and HRM indicate a robust meal response (at ∼75 min) which continues for the duration of the postprandial period analyzed. BSCM and HRM also both indicate a strong increase in motility stimulated by bisacodyl. In this patient, manual analysis identified HAPCs commencing about 10 min after bisacodyl was introduced into the colon. The HAPCs occurred in two distinct bouts over a ∼25-minute span (t∼175 - 200 min) with a ∼6 minute period of quiescence in between. The BSCM-HRM MI correlation coefficient was 0.71 for the meal response and 0.65 for bisacodyl, highlighting similar outcomes observed between the two measurement modes.

**Fig. 1.**
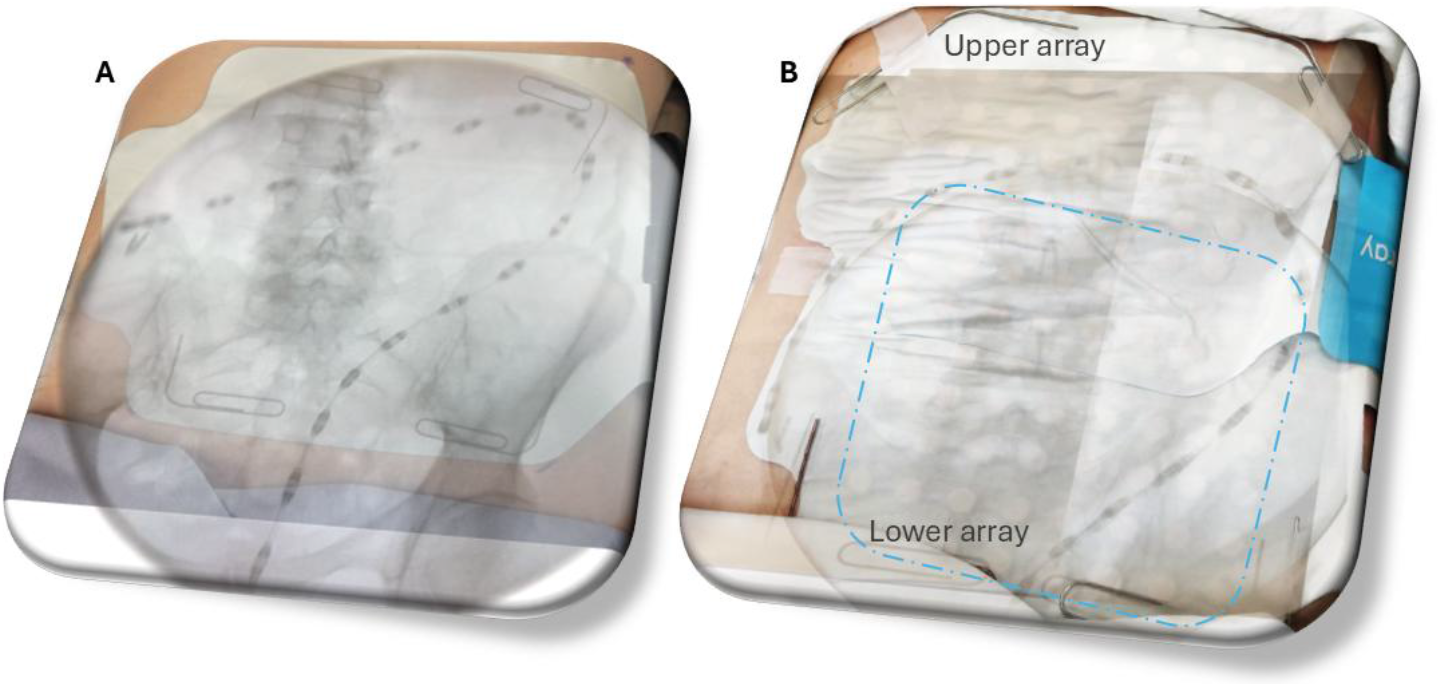
Overlaid photograph and fluoroscopy images register position of BSCM cutaneous electrode arrays and HRM intraluminal sensors. Individual HRM sensors appear as black cylindrical ‘dashes’ outlining the position and shape of the colon along the catheter’s extent. **(A)** Sufficient coverage was achieved in some subjects with a single BSCM electrode array. **(B)** Two BSCM arrays were used in patients presenting with a larger abdominal surface area. The blue rounded rectangle marks the area coverage of the lower array. The bottom rows of the upper array are not in electrical contact with the skin as they overlap the top of the lower array.

**Fig. 2.**
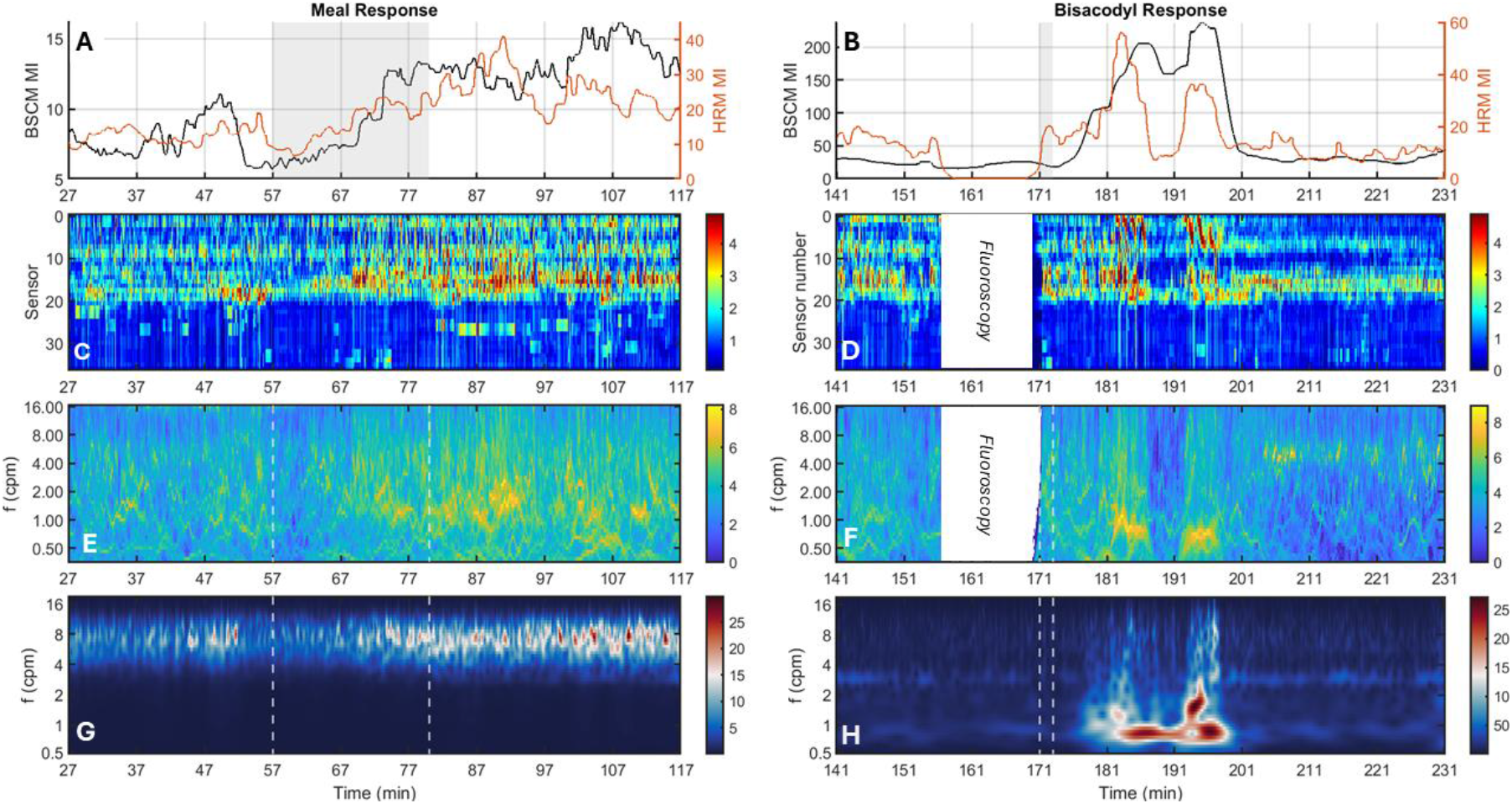
Comparison of colonic motility detected by BSCM and HRM during meal and bisacodyl response epochs. **(A** and **B)** Motility index for BSCM (black trace) versus HRM (red trace) during the meal and bisacodyl response epochs. Shaded gray rectangles indicate the timing of the respective stimulus. **(C** and **D)** HRM pressure sensor vs time 2-D visualization. Color bar indicates pressure (units of mmHg) on log2 scale. **(E** and **F)** HRM spectrograms (log2-scaled, arbitrary units) indicate dominant frequency corresponding to motility events. The HRM catheter was disconnected during fluoroscopy imaging from 156-168 min. **(G** and **H)** BSCM spectrograms. Color bar indicates amplitude in units of μV. The BSCM meal and bisacodyl responses are shown in the 4-10 cpm and 0.5–10 cpm filter bands, respectively.

As one means of assessing generalizability, we analyzed cohort-averaged MI and spectrograms. For the meal response (Fig. 3A and C), filtering BSCM signals in the 4-10 cpm frequency band yielded an optimal correlation with HRM (Pearson correlation *r* = 0.86), consistent with previous work *(11)*. For the bisacodyl response (Fig. 3B and D), filtering BSCM signals in the 0.5-10 cpm frequency band yielded the strongest correlation with HRM (*r* = 0.69).

**Fig. 3.**
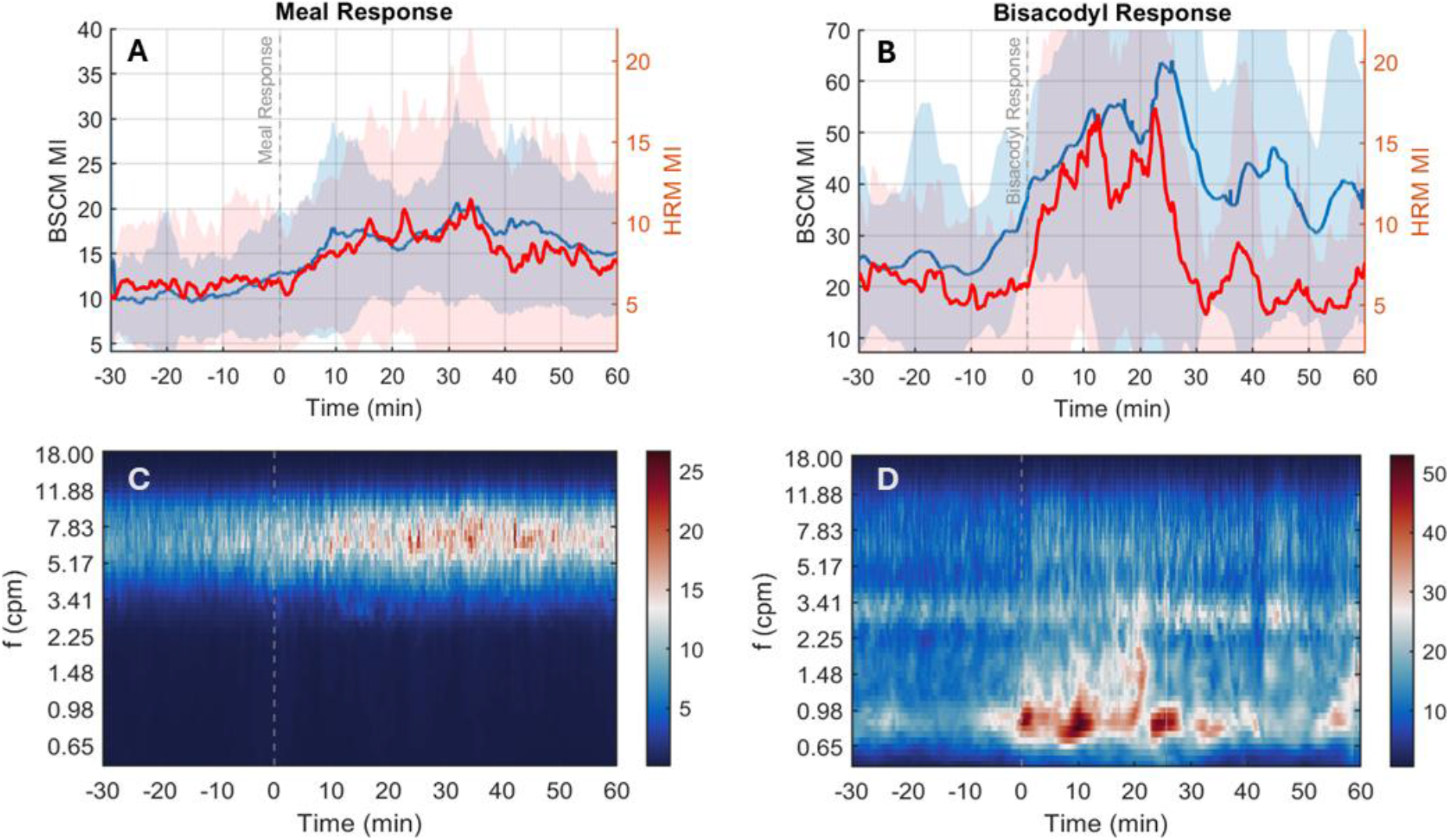
Cohort-averaged motility responses to meal and bisacodyl stimuli. (**A** and **B**): Comparison of motility index (MI) for BSCM (blue trace) vs HRM (red trace). The back shaded areas indicate intersubject variance. (**C** and **D**): BSCM spectrograms in 4-10 cpm band highlights the meal response and in 0.5-10 cpm band highlights the bisacodyl response. A large and rapid rise is observed almost immediately following the stimulus with a duration limited to about 30 min, concordant with previous observations in pediatric patients with treatment-refractory constipation (12).

### BSCM visual analysis accurately identifies bisacodyl-induced colonic motility

The level of agreement in manually marked binary classification labels (yes/no stimulus response) for HRM and BSCM bisacodyl response are summarized in Fig. 4A. The diagonal elements of the confusion matrices represent where the outcomes of HRM and BSCM agree. The off-diagonal elements represent where the labels for HRM and BSCM disagree. The 0.5 -10 cpm filter band achieved strong agreement for bisacodyl response categorization (87.5 ± 9.6%). Correspondingly, the sensitivity (*Sens*) was 84.65 ± 8.6%; the positive prediction value (*PPV*) was 100%, with notably no false positives cases possible; thus’ achieving an overall performance metric of *A*_*roc*_ = 87.5 ± 8.5% (Fig. 4B).

**Fig. 4.**
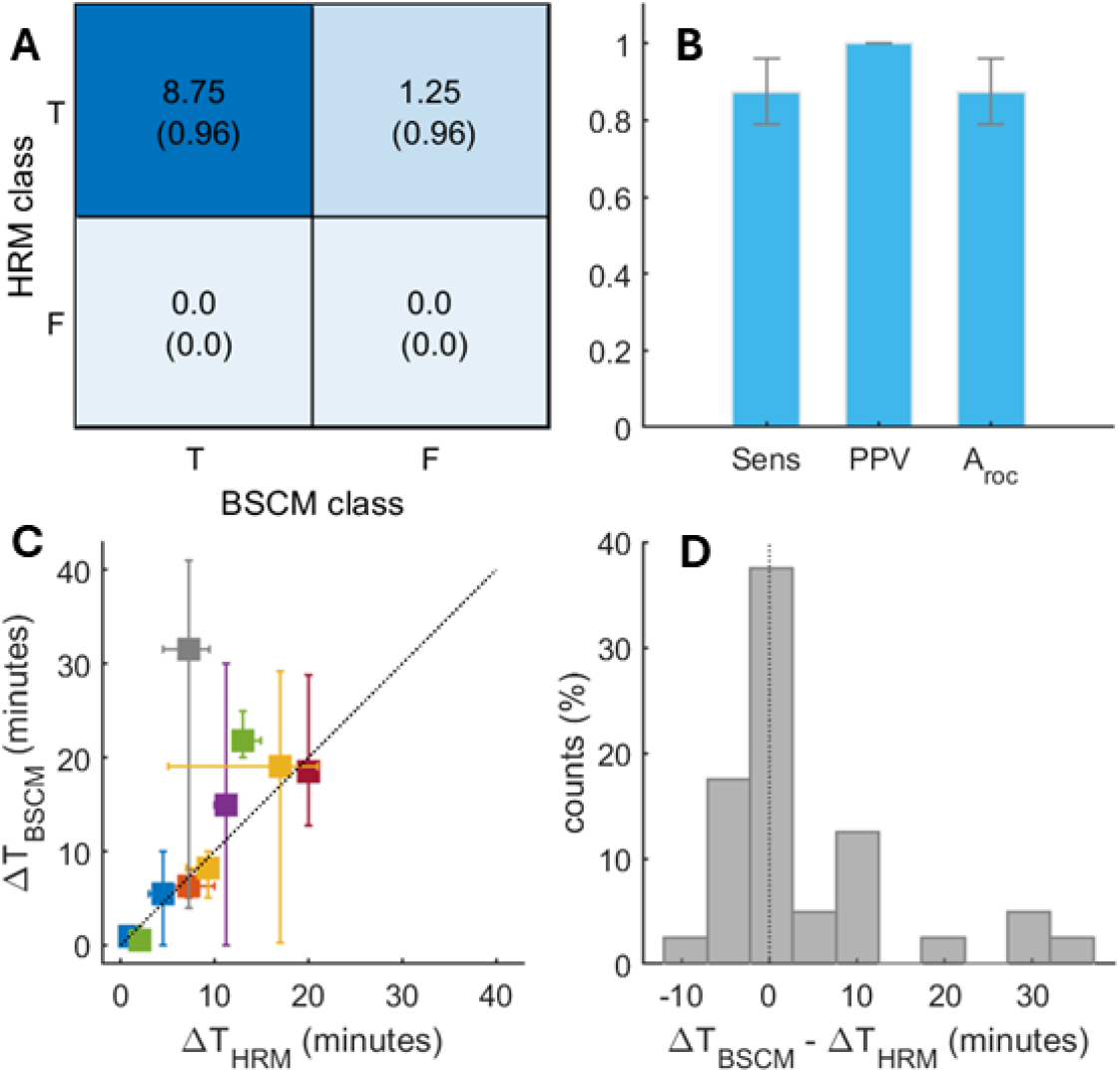
Visual diagnostic performance and temporal concordance of bisacodyl responses. **(A)** Confusion matrix for cohort-wide categorical agreement between BSCM and HRM. Printed text values indicate mean *±* (SD) across manual markings. Elements on the main diagonal represent the same outcome (true positives and true negatives), whereas off-diagonal elements indicate dissimilar outcomes (false negatives and false positives). **(B)** Performance metrics sensitivity (Sens); specificity (Spec’); and area under the ROC curve (A_roc_) summarize diagnostic performance. **(C)** BSCM vs HRM response time concordance. Each data point represents median response times and error bars extent indicates min and max values manually marked using BSCM and HRM data streams for each patient, respectively. Dotted black line (*x = y*) serves as a guide to the eye. **(D)** Distribution of differences in BSCM and HRM response times. Positive values indicate BSCM was marked as having a delayed response time relative to HRM. Dotted vertical line (value of 0) indicates the ideal outcome.

### Response Time Concordance

The response times (motility change relative to start of stimulus) marked via visual inspection HRM and BSCM streams were in good agreement (Fig 4c and d). A majority (27/34 = 79.4%) of HRM and BSCM response times differed by <10 minutes. The singular outlier with a 24.75 min discrepancy at (ΔT_HRM_, ΔT_BSCM_) = (6.75, 31.5) minutes is accounted for by a single patient case where 3 of 4 manual reviewers marked the BSCM response occurring substantially later than using HRM. On net, BSCM response times were delayed relative to HRM by only 0.5 ± 4.0 min (median ± MADM). Excluding the outlier cluster, the value of Lin’s Concordance and Pearson Correlation Coefficients were 0.79 and 0.88, respectively. The majority of marked bisacodyl response times occurred within 20 minutes of the infusion (89.7% for HRM; 71.4% for BSCM). The majority of BSCM response times were between 8.25 ± 7.25 min (median ± MADM), with a range 0.5-31 min, compared well with the time to first HAPCs event (7.3 ± 3.8 min, range 1.3 - 13.6 min) from HRM analysis.

### BSCM identifies variable symptom-motility profiles

BSCM motility-symptom associations varied across the patient cohort. Figure 5 illustrates example divergent profiles observed in two patients. In the first (Fig. 5A and B), a modest and sustained increase in the total symptom score is observed to occur synchronously with BSCM-defined meal response motility beginning at t∼80 minutes (*r* = 0.77). In the second subject (Fig. 5C and D), the total symptom score indicates a higher baseline burden which decreases during the meal response (*r* = 0.29). A large and synchronous change in symptom burden and bisacodyl-induced motility was observed in both subjects (*r* = 0.86 and 0.82, respectively). Individual symptom-BSCM motility profiles for the full cohort are provided in Supplementary Material.

**Fig. 5.**
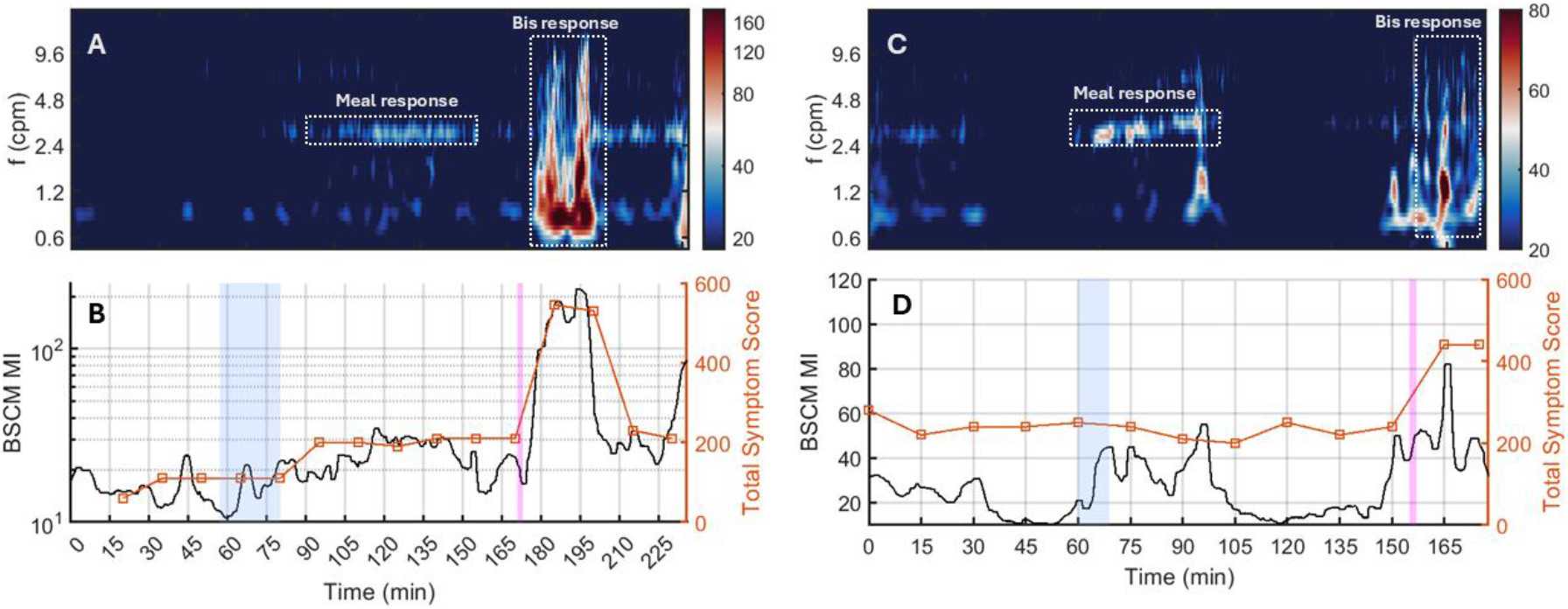
Divergent symptom-motility profiles identified from BSCM. **(A)** BSCM spectrogram and **(B)** motility index (black trace) versus lower gut symptom burden (red trace) for a patient with symptom burden increases highly correlated to motility evoked by both meal and bisacodyl. Logarithmic scaling facilitates simultaneous visualization of low and high-amplitude motility events. **(C)** BSCM spectrogram and **(D)** motility index versus low gut symptom burden for a patient with symptom burden correlated to motility induced by bisacodyl but not the meal stimulus. Meal and bisacodyl motility responses identified highlighted in spectrograms by white-dotted boxes. Color bars indicate amplitude in units of μV.

## DISCUSSION

This study presents a substantial advance in BSCM technology over earlier feasibility studies in controls *(11, 13)*. The primary outcome was validating that BSCM can accurately and reliably detect HAPCs induced by bisacodyl. Through optimized signal processing, we observed strong concordance between BSCM and gold standard HRM. In addition, BSCM was shown here to reliably detect the colonic meal response in an adult chronic constipation cohort, building on prior work in healthy controls *(11)*. BSCM also differentiated distinct symptom-motility profiles observed across this cohort (see Supplemental Materials fig. S5), demonstrating feasibility that it could be used to elucidate mechanisms involved in colonic dysfunction.

While the overall agreement between HRM and BSCM for the detection of the bisacodyl response was strong there were some discrepancies between BSCM and the manual detection of HAPC. For the binary classification task, the 5 false negatives out of 40 labels (12.5%, Fig. 4a) were accounted for by two patient cases. Both had lower signal-to-noise ratio electrical recordings, with a restricted region of electrodes with high impedance contact (>500 kΩ). One of these patients had a subpubic catheter, thus affixing the BSCM array to the abdominal surface was more challenging than normal. Therefore, it is likely that in both cases the poor correlation resulted from methodological issues associated with attaching the body surface array to skin.

The timing from ingestion of a meal or introduction of bisacodyl to the colonic motor response is of potential diagnostic importance. In our study there was a very good agreement between the manometry and BSCM bisacodyl response times. However, in one patient, manual analysis marked the BSCM response ∼25 min later than the corresponding response marked in the manometry. In this instance, the smaller peak of the true bisacodyl response is overshadowed by a second, larger peak, which was subsequently identified as the bisacodyl response. This second peak could be an artefact caused by body movement; however, no such movement was detected by the accelerometer-derived activity index or changes in impedance. Furthermore, as this peak was only observed in the lower frequency range (< 4 cpm) and did not manifest in the 4–10 cpm range, it suggests that the BSCM detected genuine gut myoelectrical activity. Bisacodyl is not reported to excite small bowel motility, indeed manometry suggests gastric and small bowel manometry becomes inhibited during bisacodyl induced HAPC, so it is unlikely this exaggerated response comes from another region of the gut *(14)*. Future work may integrate information from multiple frequency bands to more robustly identify the presence (or absence) and onset time of colonic motility patterns induced by a stimulant laxative.

Motion artefact and poor skin contact can influence the electrical signals captured by BSCM and given bisacodyl can induce cramps, abdominal discomfort and urgency, it might be argued that the large transient electrical waveforms observed during the bisacodyl were the result of external factors. However, in this study we do not consider this to be plausible based on the results of the mediation model indicating that colonic myoelectric activity was more likely to be the signal source (see Supplementary Material, fig. S9 and fig. S10). HAPC detection from the body surface was clearly validated for the first time in this study. The wideband HAPCs spectral signature observed in BCSM (Fig 3d and supplementary S1C) is consonant with the repertoire of myogenic and neurally coordinated electrical oscillations and their mechanical correlates that have been previously reported in the human colon *(15)*. The prominent ∼0.5 - 1.25 cpm spectral peak observed in BSCM is in strong accord with the ∼1-4 minute interval between successive HAPCs empirically determined via HRM, which are hypothesized to be neurally coordinated *(8, 16, 17)*. The minor peaks at 3cpm and 8cpm in BSCM are consistent with the intrinsic oscillation frequencies of Interstitial Cells of Cajal (REF). There have been prior reports showing bursts of electrical oscillations at ∼20-28 cpm underlying coordinated mechanical contractions at 3.4 - 6.3 cpm *(18)* measured in vitro on the serosa of excised colonic tissue, as well as high frequency bursts of electrical activity recorded in vivo from the mucosa with corresponding contractions at characteristic frequencies 2-3 and 10-12 cpm *(19)*. We have not attempted to identify oscillating components in this frequency range with BSCM, but do not anticipate they would be dominant components, on account of volume conduction spatiotemporal low pass filtering *(20)* as well as spatial averaging over the electrode contact area *(21)*.

There are several limitations with the present study. This study was completed in the prepared bowel and required catheter insertion under mild sedation. Although bowel cleansing has been reported to have minimal influence on the meal response, it is reported that both the frequency and amplitude of HAPC increase in the prepared bowel compared to the unprepared bowel *(22)*. Previous reports indicate that administering sedatives may attenuate colonic motility responses for several hours *(23, 24)*. However, motor patterns recorded in an unprepared colon are also seen in a prepared colon within hours of aesthesia. While the number of colonic events may differ, the overall responses to physiological and chemical stimuli remain the same.

While the relatively small patient cohort size presented in this study was sufficient to validate BSCM vs HRM, the full spectrum of possible motility responses may not be represented. Notably, all patients exhibited strong responses provoked by bisacodyl: 9 with HAPCs, 1 with high-amplitude uncoordinated (non-HAPCs) activity. Given the strong correspondence between HRM and BSCM in the present study, we hypothesize that BSCM will be robust to false event detection. A supporting preliminary result for this assessment comes from an ongoing parallel study in pediatric patients, in which BSCM correctly identified a pair of non-responders, concordant with HRM analysis *(25)*.

Whereas HRM yields detailed spatiotemporal analyses of HAPCs, such as site of origin and propagation direction and extent, BSCM source localization remains challenging, owing to geometry of the colon leading to variable source-sensor distance. Segments of colonic tissue positioned toward the anterior project more strongly onto the electrode array than those in the posterior. Modeling an active segment (ring) of colon tissue as a simple dipole whose electrical potential magnitude is proportional to 1/*d*^2^, where *d* denotes the source-sensor distance, and estimating *d* ∼ 3-12 cm based on anatomical dimensions, leads to a factor of 16 (=12^2^/3^2^) difference. The situation is further complicated by the variable orientation of the colon’s central axis relative to the body surface.

Despite current limitations relative to HRM, BSCM could potentially be utilized early in the clinical diagnostic pathway. In paediatrics, the primary indication for colonic manometry in chronic constipation is to differentiate behavioral cases (reluctance to defaecate) from intrinsic colonic dysmotility. A normal study provides visual evidence of increased postprandial motility, as well as bisacodyl-induced HAPCs propagating into the rectosigmoid junction *(24)*. A study of 165 constipated children revealed that ∼93% met the bisacodyl motility increase criterion, hence BSCM implemented at an earlier stage may obviate the need to perform HRM in a large fraction of a similar patient group. While BSCM is a promising technique that could open more effective diagnostic pathways for both paediatric and adult patients, further enhancements are needed to track HAPC propagation into the rectosigmoid junction. Ultimately, a larger cohort study is required to fully evaluate its potential for guiding clinical management decisions.

In conclusion, BSCM is an emerging, non-invasive technology that demonstrates strong correlations with HRM findings while offering several distinct advantages. It is easy to implement, essentially painless, and requires neither sedation nor bowel preparation.

Furthermore, it utilizes fewer hospital resources and significantly reduces the length of stay from multiple days to just a few hours. We anticipate that BSCM will eventually guide clinical management and help elucidate the fundamental motility mechanisms underlying chronic constipation. Its application may also extend to assessing colonic function in other disorders, such as irritable bowel syndrome, Hirschsprung disease, or chronic intestinal pseudo-obstruction.

## MATERIALS AND METHODS

### Study design

This was a prospective, single-center study. Data were acquired at the University Hospital of Leuven (UZ Leuven). All experimental protocols were approved by the ethical commission (Ethische Commissie Onderzoek) of UZ Leuven. All participants gave informed written consent.

### Patient recruitment and inclusions criteria

Patients at least 18 years old with chronic constipation, who were referred to UZ Leuven for a diagnostic colonic manometry between February 2023 and May 2025, were asked to additionally undergo a simultaneous BSCM measurement. Previous investigations, current medication, and medical or surgical history were not considered for inclusion. Exclusion criteria were a history of colonic surgery and chronic use of opioids.

### Study protocol

Patients followed a low-fiber diet for three days and fasted overnight prior to the investigation, with clear fluids permitted until four hours before the procedure. To ensure adequate bowel preparation for manometry catheter insertion, patients consumed two doses of Plenvu® on the preceding day.

Under conscious sedation with up to 5 mg of midazolam and up to 50 mg of pethidine, a solid-state catheter (Laborie, Enschede, The Netherlands) with 36 sensors spaced at 3 cm was placed via colonoscopy, with the catheter tip taken as close to the cecum as possible. A loop on the catheter tip was clipped to the colonic mucosa with two hemostatic clips (Boston Scientific®). Following a recovery period (of 60 minutes) the catheter was connected to the data acquisition unit. Pressure recordings were acquired at a sampling frequency of 10 Hz.

Body surface electrical recordings were acquired using the Alimetry® data reader connected to 64-channel adhesive electrode arrays (8×8 configuration, 2 cm spacing). Prior to placement, the abdominal skin was gently exfoliated with NuPrep gel to facilitate low impedance electrical contact. For patients with a smaller abdominal area, a single array was placed over the lower abdomen centered to the left of the midline by 2-4 cm, with the lower boundary of the array typically at the waistline (Fig. 1, left panel). This placement was chosen primarily to capture electrical activity deriving from motility patterns in the descending colon and rectosigmoid junction. For 3 patients with a sufficiently large abdominal surface, a second body surface electrode was affixed, intended to capture activity from segments –typically the transverse colon—located above the lower array (Fig. 1, right panel). Placement of the upper electrode array was guided by the Alimetry App positioning algorithm, which takes into account easily identifiable anatomical landmarks: xiphoid, umbilicus, anterior superior iliac spine, abdominal circumference *(26)*. Upper array electrode rows overlapping atop the lower array (2 maximum) were not in electrical contact with the abdominal skin surface (Fig. 1, right panel). BSCM electrical signals were acquired at 4 Hz.

A standard photograph of the array on the skin surface and a fluoroscopy image of the catheter *in situ* (19.31 cGy cm^2^) were overlaid to register the position of the BSCM array(s) relative to the manometry recording sensors (Fig. 1). Paperclips serving as radio-opaque markers were attached to the edges of the electrode arrays to visualize their position in the fluoroscopy image.

After imaging, simultaneous recordings of HRM and BSCM commenced. After 1 hr of baseline measurement, patients consumed a standardized low-calorie meal (314 kCal) within 20 minutes, consisting of 4 slices of white bread, jam, butter, cheese, vanilla pudding (11.6 g protein (14.7%), 13.8 g fat (39.7%), and 35.8 g carbohydrates (45.6%)), and a glass of water. After ∼1.5 hours of postprandial measurement, stimulant laxative was administered intraluminally (through the central core of the catheter?), consisting of 10 mg of bisacodyl (2 × 5 mg Dulcolax®, dissolved in 10 mL of saline), followed by 10 mL of saline and 5 mL of air. The measurement continued for 1 hour, except for one case where the study was terminated 35 min after bisacodyl infusion due to strong defecation urge. At the end of the study, the BSCM arrays were removed and disposed of, and the manometry catheter was removed by gentle traction.

During the study patients remained in a supine position in a hospital bed and were asked to limit movement and talking. Patients were allowed to use a bedpan when they needed to go to the toilet. 3-axis accelerometer data was recorded at 1Hz and used to analyze patient motion and position off-line. Impedance measurements acquired at 4 Hz were used to assess the quality of electrical contact, as well as secondary means to assess patient motion.

Throughout the study, patients registered upper and pan-abdominal symptoms at ∼15-minute intervals over the duration of the study using a standardized and validated iPad App *(27)*. Additionally, subjects manually recorded lower abdominal symptoms on a standardized paper form, prompted at the same 15-minute intervals. Upper abdominal symptoms included epigastric pain, epigastric fullness, early satiety, epigastric burning, heartburn, and nausea. Lower abdominal symptoms included bloating, abdominal discomfort, desire to pass gas, desire to defecate, urge to defecate, and abdominal pain. Upper and lower gut symptoms were scored on a 0-100 point scale (in intervals of 10).

### HRM manual identification of HAPCs

As HAPCs are the primary motor pattern induced by bisacodyl, manual analysis of the bisacodyl response focused upon the identification of this motor pattern. Data were exported from the Laborie software and opened in PlotHRM *(6)*. HAPCs were defined as anterograde (aboral), contractions with a minimum 12 cm propagation length (5 sensors) with at least 3 sensors recording a peak amplitude ≥ 100 mmHg *(4)*. For each study we characterized the time from bisacodyl infusion to the first HAPC, the number of HAPC, and the time interval between HAPC and the duration of the response; time from HAPC to the last. Many previous studies have shown a lack of HAPC prior to an after meal in short duration studies after a bowel preparation and therefore the meal and therefore manual HAPC analysis was restricted to the bisacodyl response.

### BSCM and HRM comparison

We analyzed epochs of data 30 min prior to and 60 min following the start of the meal and bisacodyl stimuli, respectively (except for the single case where the post-bisacodyl recording was limited to 35 min). The level of concordance between HRM and BSCM was assessed using: 1) Pearson correlation coefficients of motility indices; and 2) visual analysis and manually labeling the presence (or absence) of a colonic response to the stimulus. As prior work describing detection of HAPCs using body surface electrical recordings to our knowledge does not exist, we used a range of BSCM signal pre-processing parameters—primarily adjusting digital bandpass filter cutoff limits—to optimize detection of bisacodyl responses (Supplemental Information, see figs. S2 and S3).

### Detection of meal response motility from BSCM recordings

Body surface colonic mapping (BSCM) electrical signals were processed to identify meal response motility, predominantly cyclic motor patterns *(6)*. Details of the automated signal processing methods for identifying meal response have been previously described *(11, 13)*. Briefly, the four main steps are as follows: 1) remove wandering baseline with a moving median filter; 2) attenuate out-of-band components with a digital bandpass filter; reduce short time-scale; 3) apply a temporal Wiener filter to attenuate large-amplitude transients which can result from motion artifact; and 4) further attenuate remaining high amplitude waveforms via soft-thresholding with an upper threshold set to 500 μV.

### Detection of bisacodyl-induced high amplitude contractions from BSCM recordings

The meal-response signal processing pipeline was modified to identify the bisacodyl response using BSCM, which is anticipated to manifest with much higher amplitude signals on the body surface. Specifically, the soft thresholding step was removed from the signal processing pipeline, to avoid attenuating short-lived (order of ∼1 minute), large amplitude (>200 μV) waveforms, which are likely to be the electrical correlates of high-amplitude colonic motility patterns (primarily HAPCs). To optimize BSCM detection of high-amplitude waveforms, we quantified performance using a range of bandpass filter cutoff frequencies including: 0.12 -12, 0.2–2.2, 0.6 - 6.0, 0.5–10, 4-10 cycles per minute (cpm). The lower bound was motivated by previous reports *(12)* and HRM manual analysis in this patient cohort quantifying the interval between successive HAPCs was densely distributed in the range of 1-4 minutes—equivalently 0.25 - 1.0 cpm. The rationale for upper cutoff limits was to pass higher frequency components of the signal expected to be a frequency range of 2-12 cpm band *(15, 19)* while also mitigating potential source interference respiration (∼12-20 breaths/minute in adults). Finally, we computed the BSCM motility index (MI) detailed in *(11)*, which is based on summing the largest 10% of coefficients in the spectrogram at each moment in time.

### HRM automated analyses

We analyzed HRM recordings with automated methods previously described *(28–30)*. This analysis uses synchrosqueezed wavelet transform spectra with application of the ‘Mesaclip’ algorithm to remove harmonics, thus emphasizing the fundamental frequency (i.e. the inverse of time intervals) at which motility events occur. Data were pre-processed to remove baseline drift and all synchronous pressure increases spanning all pressure sensors. Pressures below 1mmHg were clamped at the threshold value, and log2-transformed so that high-amplitude events would not overpower potentially significant low-amplitude oscillations, which is important for proper visual interpretation. The HRM motility index (MI) was defined as the sum of the spectrogram coefficients at each moment in time. In computing the MI, we included only sensors positioned directly under the BSCM array, as judged from careful inspection of overlaid fluoroscopy images and overhead photographs (e.g., Fig. 1).

### BSCM and HRM motility index correlation

BSCM and HRM data files were time-synchronized. We utilized their respective MI to compare mechanical activity to electrical signals. Specifically, we quantified the level of agreement between these measurement modes with the Pearson correlation coefficient. In addition, we computed the maximum cross-correlation value (‘Max Corr’) between the BSCM and HRM MI, limiting the time lag to ±5 min. This was a heuristic metric to account for known imprecision in manually recorded time stamps, typically recounted to be on the order of a few minutes, that naturally arise in a busy clinical environment.

### Visual analysis and manual categorization of meal and bisacodyl responses

Experienced researchers (*n=*4) reviewed a series of figures showing outputs from HRM and BSCM automated analyses (e.g., see Fig. 3) and asked to identify whether a meal response and whether bisacodyl response occurred in each case. Binary classification labels (yes/no) were assigned for each case. This simple assessment was intended to correspond to visual identification performed in the clinical environment. BSCM manual review was limited to the best three performing parameter combinations per Pearson correlation analysis (passbands of 0.12 - 12, 0.5 - 10, and 4-10 cpm). In sum, 80 panels were reviewed and labeled: 10 subjects x 2 stimuli x (1 HRM panel + 3 BSCM panels). In cases where a response was judged to occur, reviewers were asked to notate the time at which the motility response began.

Confusion matrices were computed to summarize the level of agreement between HRM and BSCM manual reviewer binary classification labels. The diagonal elements indicate cases where HRM and BSCM yielded the same result (true positive *TP* and true negatives *TN*); the off-diagonal elements indicate cases where the labels disagreed (false positives *FP* and false negatives *FN*). To further quantify the level of agreement, we computed the sensitivity *Sens* = *TP*/(*TP* + *FN*), positive predictive value *PPV* = *TP*/(*TP* + *FP*), and the overall performance metric as the area under the receiver operating characteristic *A*_*roc*_ = *Sens* × *PPV*.

### BSCM symptom-motility relationships

To compare the BSCM MI to symptom scores, we averaged the BSCM MI in a 10-minute block centered on each symptom score time point. This was done to account for the lower time resolution of symptom reporting and the uncertainty of the exact timing at which the symptom(s) were experienced and recorded relative to BSCM signals which were continuously monitored.

### Motion and impedance change artifact signal components

Given bisacodyl infusion is likely to cause pain and cramping, we anticipated that patients may change posture or press their arms over the abdominal area where they feel pain, thus pressing against the electrode array. These actions may generate motion or impedance change artifacts, respectively. We utilized a causal linear mediation causal model to quantify their contribution to recorded waveforms relative to internal colonic electrical sources (see Supplemental Information for implementation details).

### Statistical Analysis

Sample statistics are reported in most cases as the mean ± SD. The median ± median of the absolute deviation (denoted MADM) was used as robust metrics to summarize data known to include outliers or skewed distributions, including BSCM-HRM manually identified response times as well as manually identified HAPCs event intervals. The MADM is related to the via SD ∼ 1.48 x MADM, with the MADM relatively insensitive to bias by extremum values.

## Data Availability

The data generated in this work will be made available upon reasonable request. Requests should be made to the corresponding author (JCE).

## List of Supplementary Materials

Fig S1 to S9

Tables S1 and S2

References (*32*–*33*)

## Acknowledgments

We thank Gabe Schamberg for helpful discussions on mediation model analysis.

## Author contributions

Conceptualization: JT, JCE, GOG

Methodology: AV, PD, JCE

Investigation: AV

Formal Analysis: AV, JCE.

Visualization: JCE

Software: JCE

Funding acquisition: JT, GOG

Supervision: JT

Writing – original draft: PD, JCE, AV Writing – review & editing: PD, JCE, GOG

## Data and materials availability

The data generated in this work will be made available upon reasonable request. Requests should be made to the corresponding author.

