## Supplemental Materials for "Body surface colonic mapping detects meal and bisacodyl-induced colonic motility in patients with chronic constipation"

**Electrical correlates of mechanical activity**

Figure S1 illustrates simultaneous HRM and BSCM recordings centered on the bisacodyl response, highlighting the electrical correlates of discrete propagating (pressure) waves of contraction. Time domain signals highlight the tight time synchrony between HAPCs observed in HRM recordings (fig. S1A) and the corresponding large amplitude (~1.5 mV peak-to-peak) oscillations observed in BSCM recordings (fig S1B). Whereas the HRM recording indicates two distinct bouts of HAPCs with ~6 min pause between contractile activity, BSCM indicates continued oscillations with a modestly decreased amplitude during the pause. The power spectral density indicates prominent peaks concentrated in the ~0.7 – 1.2 cpm frequency band for both (fig. S1C). BSCM also indicates smaller frequency peaks at ~3 and 6-9 cpm, consistent with previous findings using HRM *(15, 31)*. These findings are consistent with the hypothesis that HAPCs are generated when underlying myoelectrical oscillations summate via coordinating extrinsic nervous input *(32, 33)*.

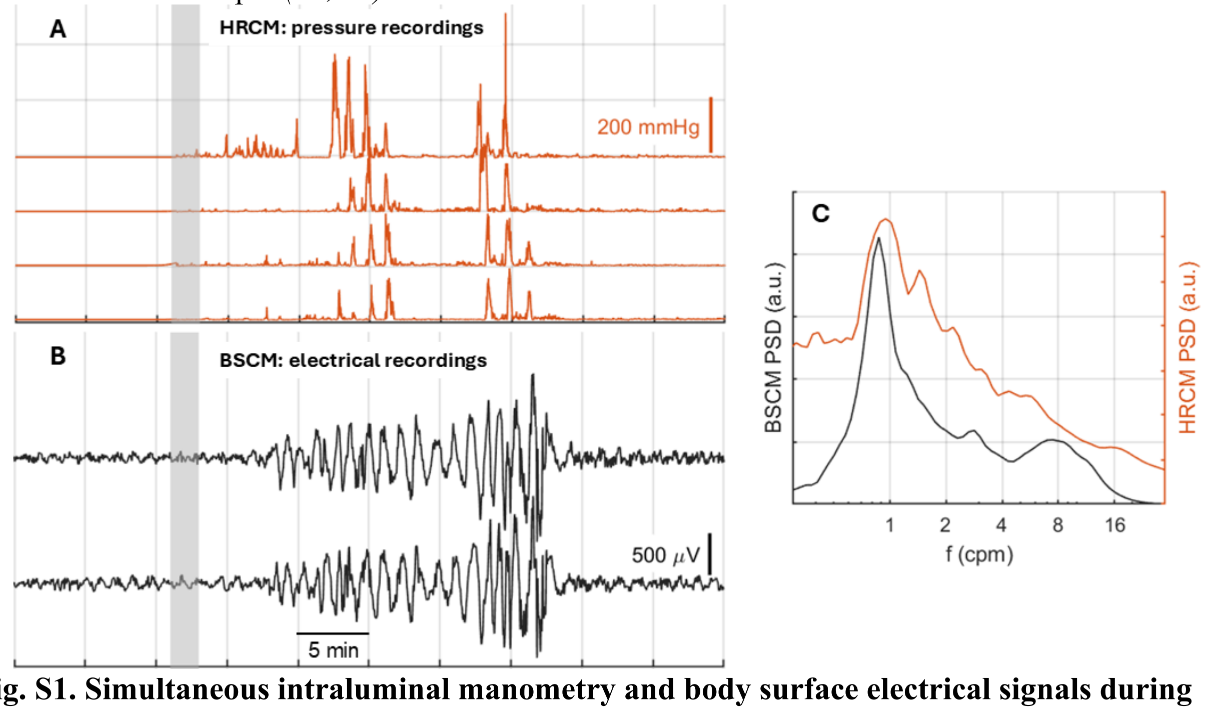


**fig. S1. Simultaneous intraluminal manometry and body surface electrical signals during bisacodyl response. (A)** HRM recordings indicate two episodes of HAPCs propagating in the antegrade direction. Only 4 adjacent pressure sensors are shown for clarity. **(B)** BSCM recordings indicate large amplitude oscillations starting and stopping in synchrony with HAPCs. BSCM signals shown were bandpass filtered 0.12-12 cpm. The timing of bisacodyl delivery is indicated by the light gray rectangles. **(C)** Power spectral density for HRM (red) and BSCM (black). Note the logarithmic frequency scale. The data shown correspond to the bisacodyl response in Fig. 3.

**Optimizing BSCM signal processing parameters to identify colonic motor patterns**
To our knowledge, there is no prior report identifying the occurrence of HAPCs (bisacodyl-induced or otherwise) with body surface electrical recordings. It was therefore important to optimize the signal processing chain. In doing so, we sought to optimize BSCM parameter sets by evaluating outcomes for each individual subject across the patient cohort, using a combination of BSCM-HRM correlation (fig. S2), visual analysis of response (fig. S3), and response time concordance (fig. S4). The best three parameter sets were identified as maximizing BSCM-HRM MI correlation (fig. S2) were: 0.12 -12 cpm, 0.5 – 10 cpm, 4-10 cpm with the Wiener filter applied in each case (listed as option 'B' in filter limit labels in fig. S2). The correlation mean ± SD, (median) values were, respectively: 0.32 ± 0.32 (0.38); 0.32 ± 0.32 (0.40), 0.33+0.3 (0.38). When considering the maximum cross correlation ('Max Corr') allowing for time shifts of up to 10 minutes (fig. S2., middle panel), these values increased to: 0.46 ± 0.20 (0.50); 0.45 ± 0.20 (0.51); 0.45 ± 0.19 (0.42) respectively. While most differences between Max Corr and the correlation coefficient were modest, two subjects showed a substantial increase (fig. S2, right panel). These cases probably represent a lack of precision in notating the timestamps at which BSCM and HRM signal acquisition commenced, which may be recorded hours after the event due to the inherently busy clinical environment in which studies are implemented.

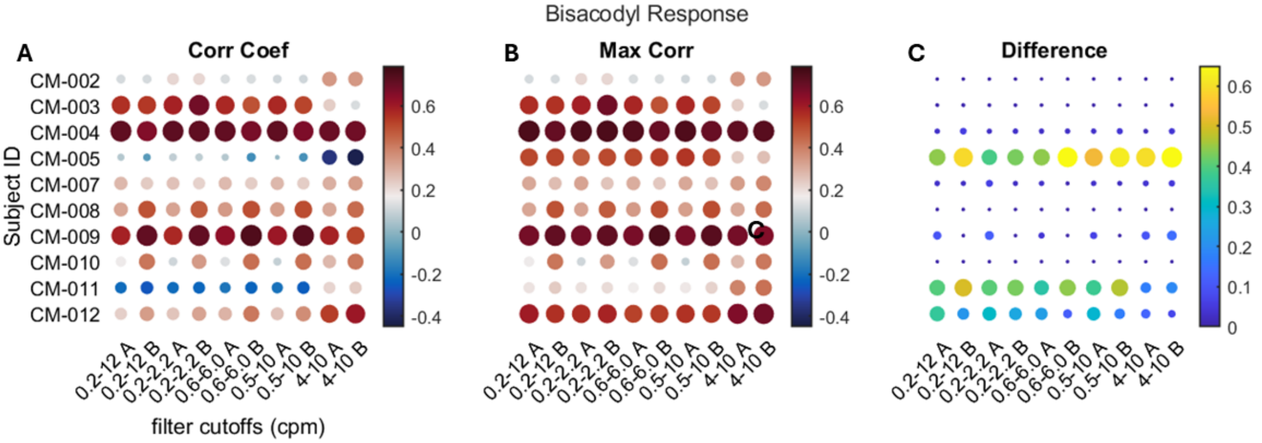


**fig. S2. BSCM-HRM MI correlation for all subjects across 10 signal processing combinations.** The parameter space included 5 filter bands (cutoffs indicated in units of cpm) × 2 Weiner filter options (A = 'off', B = 30 s analysis window and 300 s noise window). **(A)** Pearson correlation coefficients; **(B)** Maximum cross-correlation ('Max Corr') with time lag limited to ± 10 min. **(C)** Difference = MaxCorr - CorrCoeff indicates substantial increases for 2 subjects (labeled CM-005 and CM-011). Both dot size and color indicate correlation value.

Results for visual identification of bisacodyl responses (fig. S3) indicated that the 0.5-10 cpm frequency band was most effective, with a modest but noticeable decrease in performance for the 0.12-12 or 4-10 cpm frequency band. Time concordance results for bisacodyl responses (fig. S4, bottom row) indicate highly similar outcomes for the 0.2-0.12 and 0.5 – 10 cpm bands. The apparent improvement in the 4-10 cpm isn't a real gain in performance, rather the outlier cluster being 'erased' because the visual analysis labeling (erroneously) indicated 'no response', hence the points are not plotted. Recalling the near negligible differences noted between the frequency bands for the correlation-based results, we characterized 0.5-10cpm as the optimal frequency band for detection of HAPCs by BSCM.

The optimality of the 0.5-10 cpm band may be explained by the fact that successive HAPCs occur at intervals of ~1-4 min, equivalently 0.25 - 1 cpm. Hence, the lower cutoff frequency is better suited to capturing the longer time scale events. Higher frequency components, likely generated by electrical oscillations that summate into HAPCs under neurally mediated control, have been reported in the range of 8-12 cpm *(15)*. The 10 cpm upper limit is therefore a suitable compromise between passing them while attenuating possible interference sources of respiration (~10-20 cpm) and small bowel (~6 in the ileum to 12 cpm in the duodenum).

Previous work has reported ~23-28 cpm electrical oscillations correlated to longer-time scale mechanical contractions *(18, 34)*. These high frequency electrical components were measured using suction electrodes or sucrose gap recordings made from colon muscle strips in vitro. We have not attempted to identify oscillating components in this frequency range with BSCM, but do not anticipate they would be dominant components, on account of volume conduction spatiotemporal low pass filtering *(20)* as well as spatial averaging over the electrode contact area *(21)*. Nonetheless, a future study should more fully address and more completely define the full colonic signal power spectrum measured on the body surface.

**Expert visual analysis for the 3 best frequency bands**

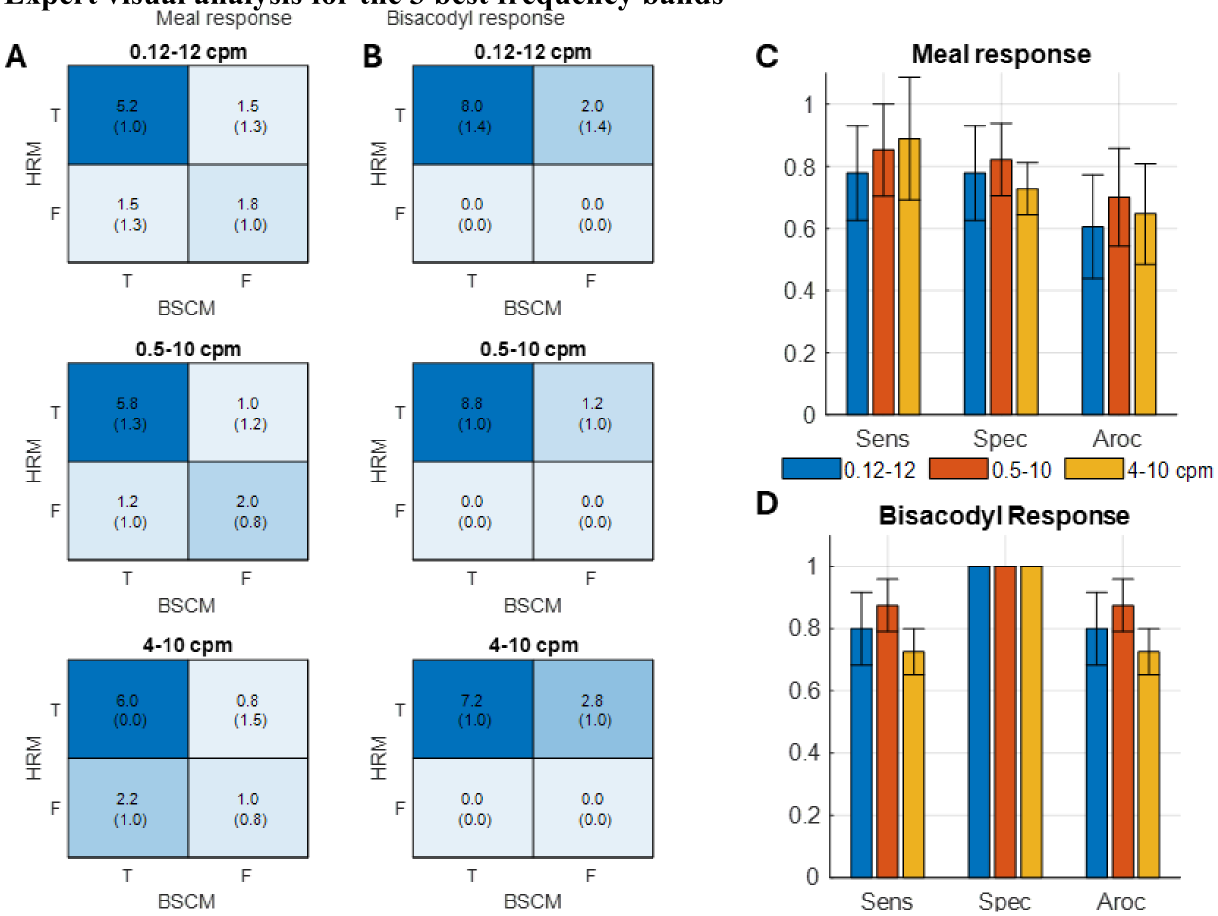


**fig. S3. Confusion matrices and performance metrics summarizing level of agreement between HRM vs BSCM.** Cohort-wide ratings are averaged across expert visual analysis manual marking (binary categorization) outcomes. (**A** and **B**) Confusion matrix for each frequency band (in rows) for meal response (left column) and bisacodyl response (right column). Text values denote mean ± (std). The main diagonal elements indicate true positives and true negatives. Off diagonal elements represent false negatives and false positives. For bisacodyl responses, the 0.5 - 10 cpm band was optimal, achieving BSCM-HRM ensemble averaged agreement in 8.75/10 cases (87.5%) on average. (**C** and **D**) Performance metric (sensitivity = Sens; specificity = 'Spec' and Aroc) summary for meal and bisacodyl responses. The 0.5-10cpm band was observed to have the maximal outcome for the bisacodyl response. The wider filter band of 0.12 - 12 cpm also performed well (Aroc = 80 ± 12%) while the narrow and higher 4-10 cpm band saw performance sag modestly to Aroc = 72.5 ± 7.5%.

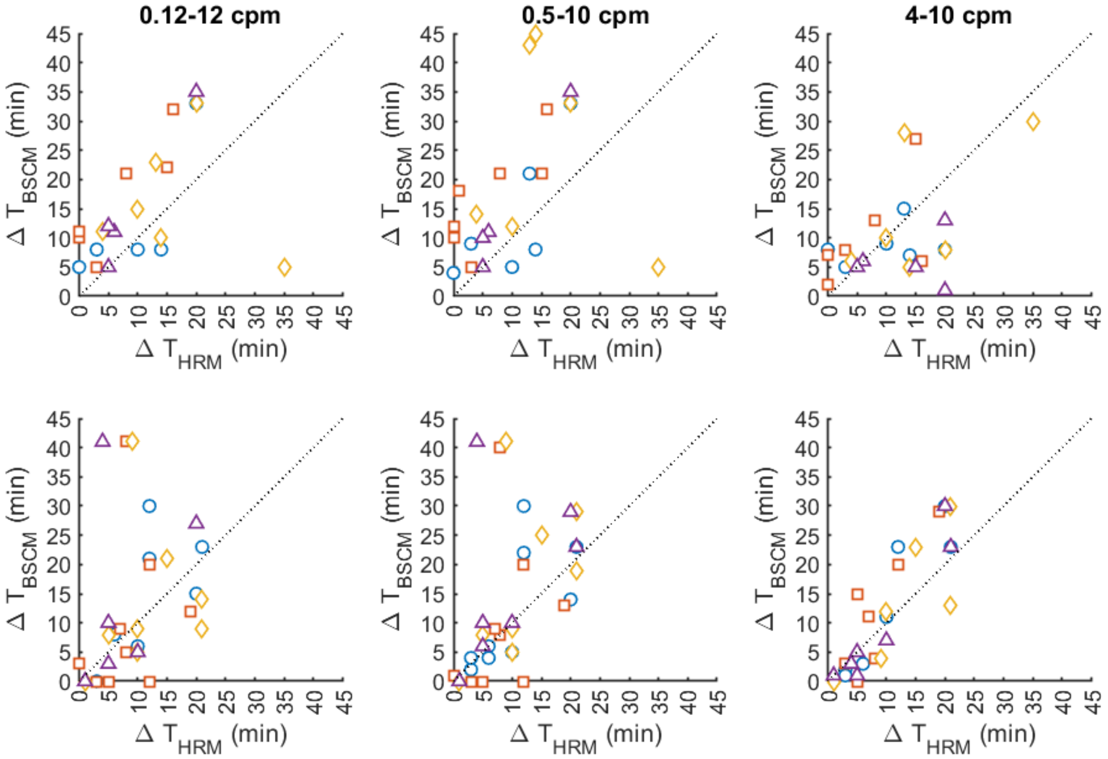


**fig. S4. BSCM vs HRM response time concordance**. a,b: meal and bisacodyl response times manually marked using BSCM and HRM data streams. Plot styles (color and shape) are unique for each of 4 manual reviewer ratings. Dotted black line x = y serves as a guide to the eye. c,d: histograms showing difference in response times marked using BSCM and HRM for meal and bisacodyl responses, respectively. Positive values indicate BSCM was marked as having a later response to a stimulus compared to HRM.

**Symptom-Motility Correlations: Full cohort**

BSCM MI-symptom relationships varied across subjects and among symptoms, with some common themes emerging. For example, most subjects reported strong bouts of discomfort and urge during the bisacodyl response synchronously to the occurrence of HAPCs (fig. S5C and F). In contrast, symptom-motility correlations were more heterogeneous during meal epochs—i.e., bloating increased during the meal response epoch evoke symptoms correlated with rises in

BSCM MI in some subjects (e.g. ‘bloating for the subject in fig. s5C), but not in others (e.g. fig. s5F). Some patients exhibited universally high symptom-motility correlations (e.g., see fig. S5H, subject 04 and fig. S5A-C) while others showed low correlation across most or all symptoms (subject 07 in fig. S5H and fig. S5B-F).

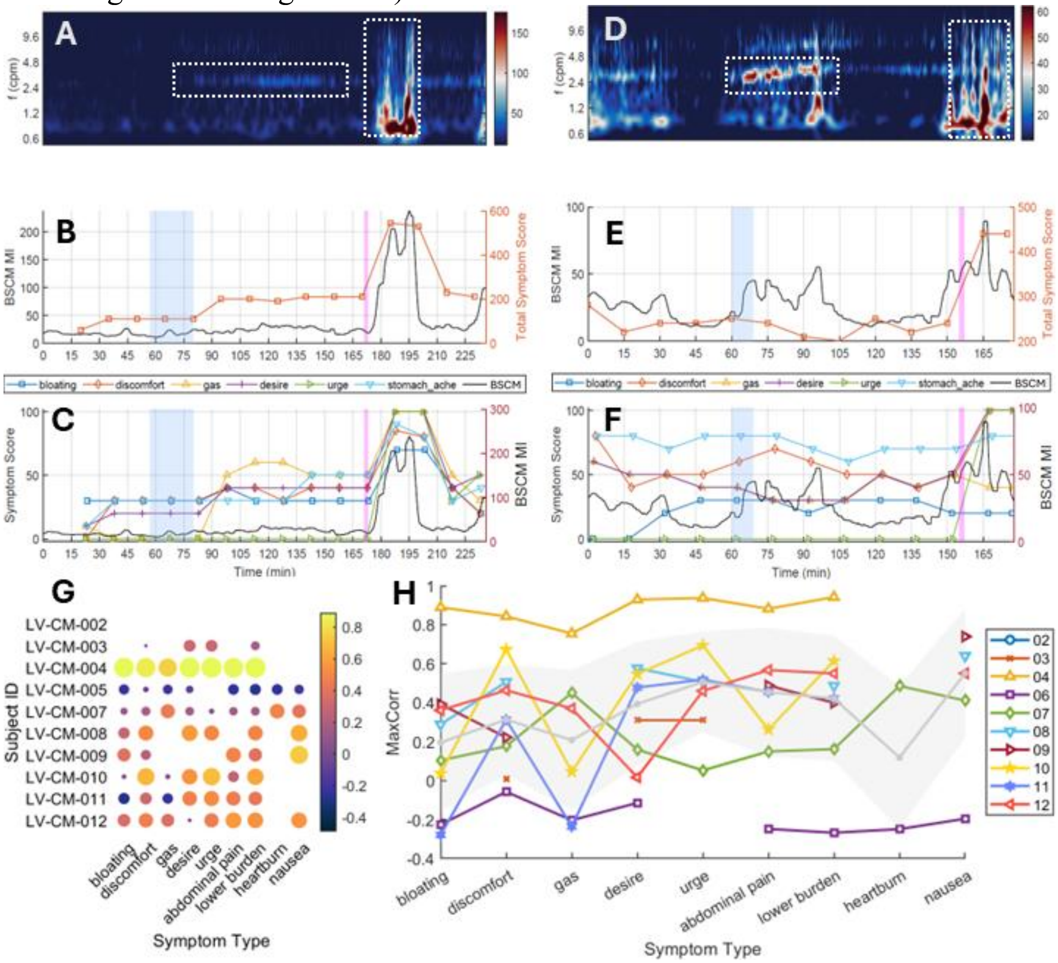


**fig. S5. Symptom-motility correlation across the study cohort. (A-C) and (D-F)** illustrates contrasting relationships for BSCM-defined motility and all lower gut symptom scores in two example patients, respectively. (**G and H**) Pearson correlation coefficients across the patient cohort and all lower gut symptoms displayed as bubble plot and line-and-scatter format. The gray data series in **(G)** represents the cohort median with backshading indicating the standard deviation.

**Analysis of meal and bisacodyl epochs**

The most pronounced increase in symptoms was observed following administration of bisacodyl, which in several cases would swamp smaller changes in motility and symptoms during the meal response period. We therefore analyzed symptom-motility relationships for meal and bisacodyl epochs separately.

**Meal response (4-10 cpm BSCM frequency band)**

Symptom-motility correlation values during the meal response epoch (fig. S6) illustrate the spectrum observed across the cohort, with cohort-wide statistics summarized in table S1. Some patients exhibited strong relationships for most symptoms (e.g. subject LV-CM-004); some showed moderate correlation across most symptoms (e.g. subject LV-CM-012); some were more specific (e.g. LV-CM-010); while for other patients there was weak or no correlation (e.g. subjects LV-CM-003 or LV-CM-009).

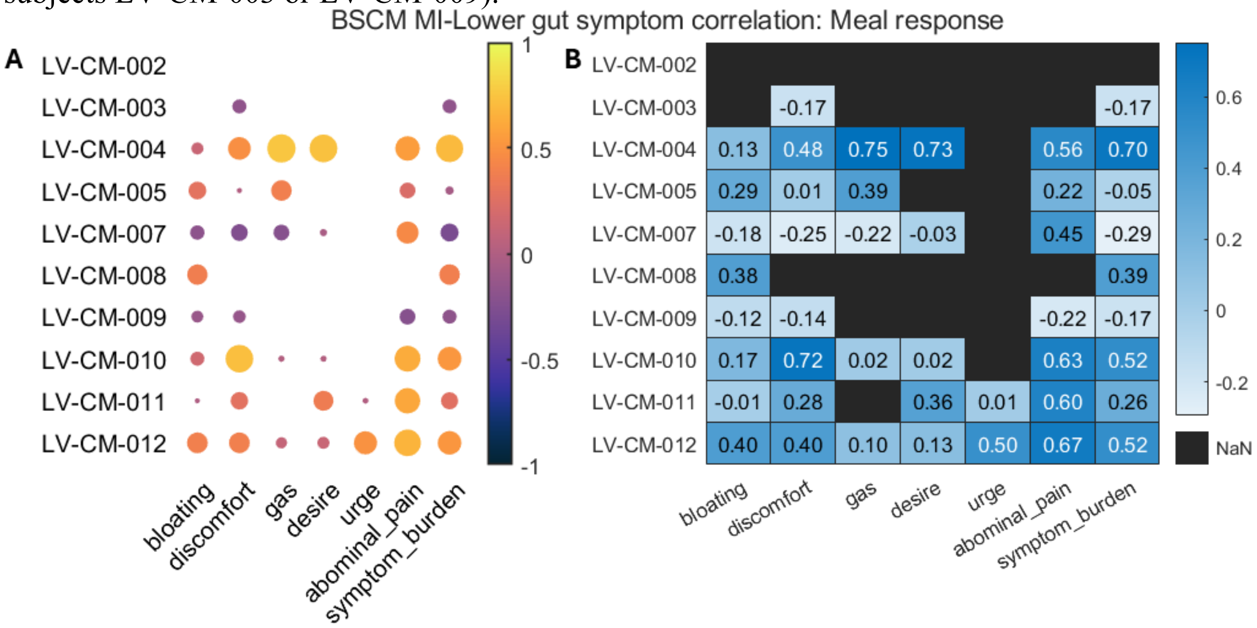


**fig. S6. Meal response symptom-motility correlations. (A)** Correlation coefficient 'bubble plot' radius and color indicates Pearson coefficient values. **(B)** Heatmap with printed text indicating Pearson correlation coefficient.

| | Bloating | Discomfort | Gas | Desire | Urge | Pain | Symptom burden |
|---|---|---|---|---|---|---|---|
| **median** | 0.39 | 0.27 | 0.31 | 0.04 | 0.30 | 0.25 | 0.29 |
| **std** | 0.26 | 0.33 | 0.32 | 0.33 | 0.02 | 0.19 | 0.29 |
| **mean** | 0.28 | 0.28 | 0.30 | 0.18 | 0.30 | 0.23 | 0.27 |

**table S1. Symptom-motility correlations for meal responses.**

### Bisacodyl Response (0.5 - 10 cpm BSCM frequency band)

Symptom-motility correlation values (fig. S7) illustrate that most patients exhibited high correlation between symptoms and BSCM-defined motility. Interestingly, the lowest correlation occurred in LV-CM-009, the singular case of a non-HAPCs bisacodyl response. LV-CM-002 shows no correlation either, however this is the singular false negative case, where BSCM was unable to identify a post-bisacodyl response, likely owing to uncharacteristically noisy (low SNR) signal quality. Cohort-wide statistics are summarized in table S2. In general, the symptom-motility correlation was high across most of the cohort while bouts of HAPCs were known to occur.

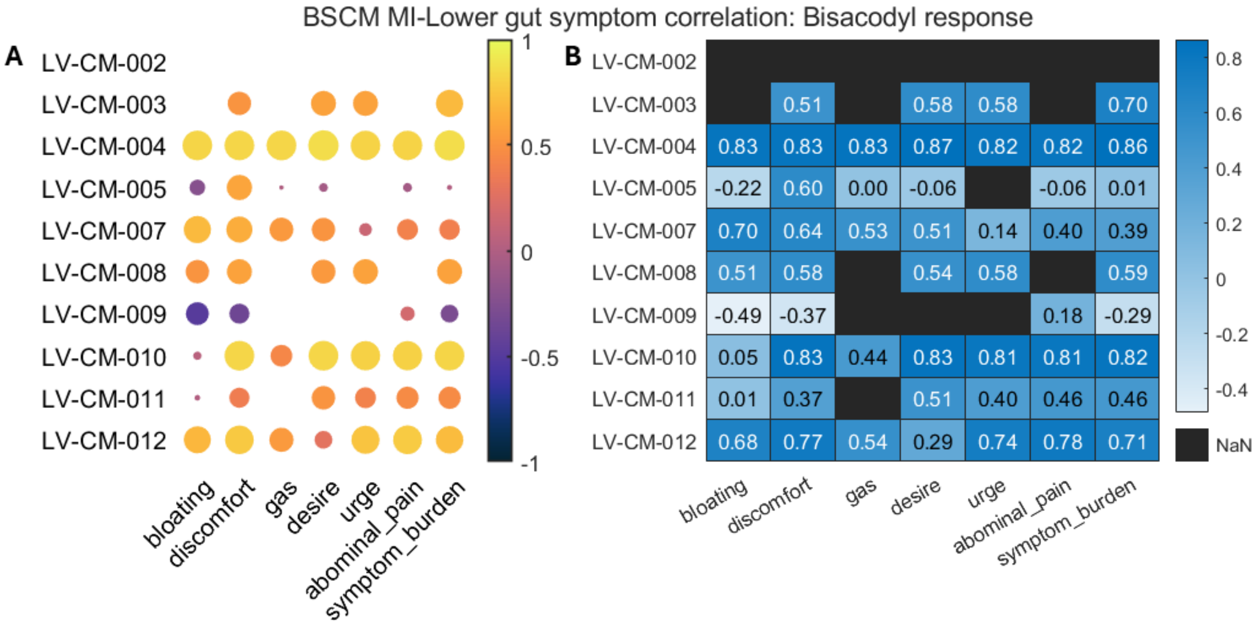


**fig. S7. Symptom-motility correlations during for the bisacodyl response epoch.** Format is the same as Fig S6.

| | Bloating | Discomfort | Gas | Desire | Urge | Pain | Symptom burden |
|---|---|---|---|---|---|---|---|
| **median** | 0.28 | 0.60 | 0.53 | 0.53 | 0.58 | 0.46 | 0.59 |
| **std** | 0.45 | 0.35 | 0.27 | 0.28 | 0.23 | 0.32 | 0.36 |
| **mean** | | 0.26 | 0.53 | 0.47 | 0.51 | 0.58 | 0.48 |

**table S2. Symptom-motility correlations for meal responses.**

**Causal pathway mediation model quantifies potential contribution of artifact sources**

We utilized a causal linear mediation model to quantify the contribution of motion artifact and or impedance change to BSCM recorded waveforms (indirect effect) relative to internal colonic electrical sources (direct effect) *(35)*. The mediation model applied to the BSCM context is indicated in fig. S8. The direct pathway indicates BSCM electrical waveforms result directly from colonic myoelectric activity, quantified by linear regression coefficient *c'*. The indirect pathway implies BSCM signals results from colonic motility which are mediated by motion artifacts or impedance changes that may manifest as large amplitude electrical waveforms. The effect of the indirect pathway is quantified by the product of coefficients ab. When *c' > ab*, we can be more confident that large electrical signals measured by the BSCM array results directly from colonic myoelectric activity rather than artifact sources. We implemented a bootstrapped (n=10000) mediation model with BSCM MI and artifact signal - either the activity index (derived from 3-axis accelerometer readings: $A.I. = \sqrt{a_x^2 + a_y^2 + a_z^2}$ or impedance change (dZ/dt in units of kΩ/s) with inputs scaled 0 to 1.

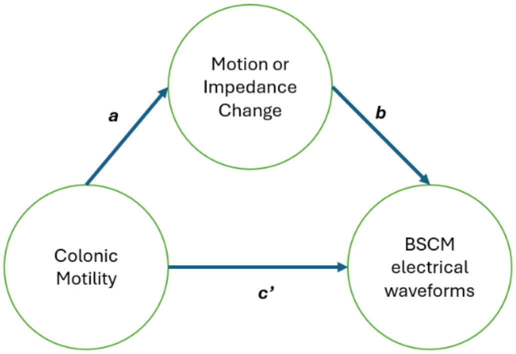


**fig. S8. BSCM mediation model.** Direct effect pathway is quantified by the coefficient *c'*. The indirect effect is quantified by the product *ab*.

Figure S9 shows the values of direct and indirect coefficient for *ab* and direct pathway *c'* for motion artifact (activity index, top row) as the mediator and impedance change as the mediator (bottom row), respectively. We analyzed causal pathways in the 3 top-performing frequency bands (in columns), identified per BSCM-HRM MI correlation analysis.

Figure S10 shows the difference *c'* > *ab*, respectively. Values greater than 0 imply a stronger direct causal pathway. Values < 0 indicate a stronger indirect pathway. Error bars indicate 5-95% confidence intervals.

For the identified optimal frequency band (0.5-10cpm), the mediation model direct effect coefficients (0.35 ± 0.33; median 0.33) were an order of magnitude larger than the indirect effect (0.02 ± 0.005, median 0.05), indicating that the large amplitude waveforms were much more likely to be generated by colonic myoelectric activity.

It is interesting to note that the causal pathways are considerably stronger for the 4-10 cpm frequency band across the cohort compared to the other frequency bands (fig. S10)

It is also interesting to note that the indirect pathways assessed with activity index (patient motion) as the mediator are unambiguous in showing a stronger direct effect. This contrasts with impedance change as the mediator. For the two frequency bands including content < 4 cpm, there are some cases where the indirect effect appears to be larger than the direct effect (subjects CM-004 and 008). The overall difference *c'*- *ab* was still positive valued on net: 0.20 ± 0.34; median 0.08. Impedance changes exhibited also higher correlation with BSCM signals during HAPCs epochs. Pearson correlation was 0.38 ± 0.30; median 0.43

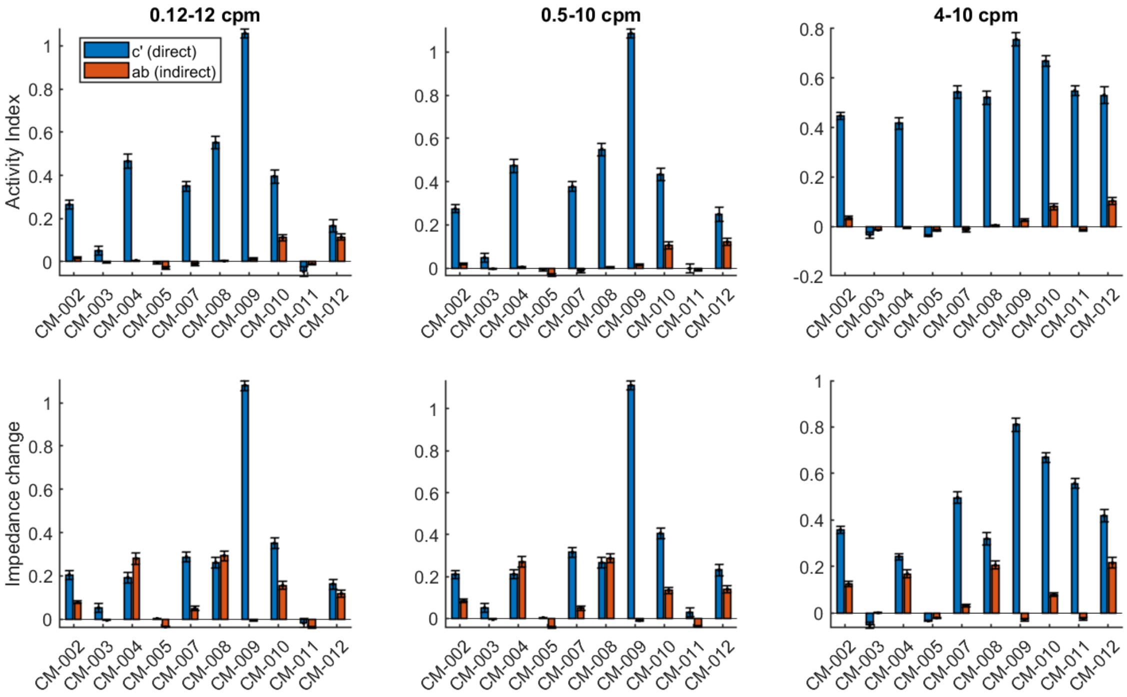


**fig. S9. Direct (blue bars) and indirect effect (red bars) coefficient values** with activity index as the mediator (top row) and with impedance change as the mediator (bottom row). The top 3 performing filter bands are arranged in rows. Error bars indicate 5-95% confidence intervals resulting from bootstrapping analysis.

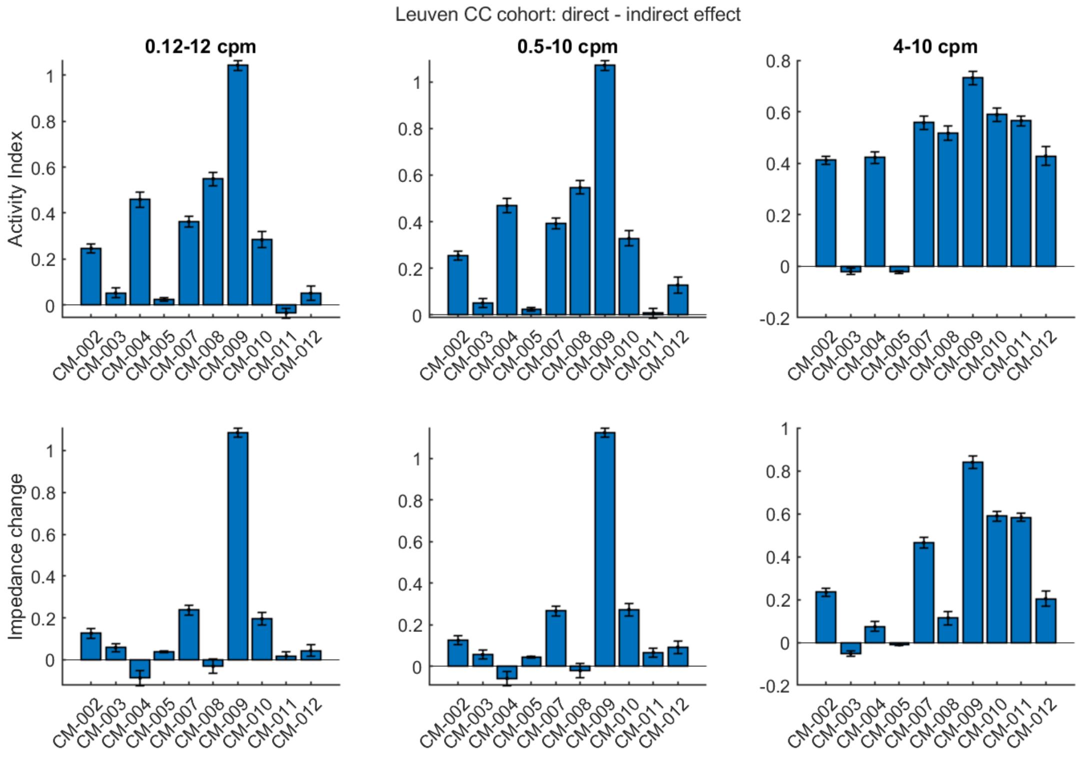


**fig. S10. The difference in direct vs indirect effects *c'- ab*.** Positive values indicate the direct effect pathway is stronger than the indirect pathway.
